# Polygenic risk prediction and *SNCA* haplotype analysis in a Latino Parkinson’s disease cohort

**DOI:** 10.1101/2021.11.09.21265082

**Authors:** Douglas Loesch, Andrea R. V. R. Horimoto, Elif Irem Sarihan, Miguel Inca-Martinez, Emily Mason, Mario Cornejo-Olivas, Luis Torres, Pilar Mazzetti, Carlos Cosentino, Elison Sarapura-Castro, Andrea Rivera-Valdivia, Angel C. Medina, Elena Dieguez, Victor Raggio, Andres Lescano, Vitor Tumas, Vanderci Borges, Henrique B. Ferraz, Carlos R. Rieder, Artur Schumacher-Schuh, Bruno L. Santos-Lobato, Carlos Velez-Pardo, Marlene Jimenez-Del-Rio, Francisco Lopera, Sonia Moreno, Pedro Chana-Cuevas, William Fernandez, Gonzalo Arboleda, Humberto Arboleda, Carlos E. Arboleda-Bustos, Dora Yearout, Cyrus P. Zabetian, International Parkinson Disease Genomics Consortium (IPDGC), Timothy A. Thornton, Ignacio F. Mata, Timothy D. O’Connor, on behalf of the Latin American Research Consortium on the Genetics of Parkinson’s Disease (LARGE-PD)

**Author notes:** Corresponding authors: Timothy D. O’Connor, University of Maryland School of Medicine, 670 W. Baltimore St., Baltimore, MD, 21201, USA. E-mail addresses, Ignacio F. Mata, Lerner Research Institute R4-006, Cleveland Clinic Foundation, 9500 Euclid Ave., Cleveland, OH, 44195, USA. E-mail addresses. Data for this manuscript was generated while IFM was affiliated at the VA Puget Sound and the University of Washington.

## Abstract

**Background:** Large-scale Parkinson’s disease (PD) genome-wide association studies (GWAS) and meta-analyses have, until recently, only been conducted on subjects with European-ancestry. Consequently, polygenic risk scores (PRS) constructed using PD GWAS data are likely to be less predictive when applied to non-European cohorts.

**Methods:** Using GWAS data from Nalls et al. 2019, we constructed a PD PRS for a Latino PD cohort (LARGE-PD) and tested it for association with PD status. We validated the PRS performance through testing the PD PRS in an independent cohort of Latino PD patients and by repeating the PRS analysis in LARGE-PD with the addition of 440 external Peruvian controls. To explore the global distribution of PD PRS, we utilized 1000 Genomes Project (1KGP) and Peruvian Genome Project (PGP) data to estimate PD risk allele frequencies. We also tested *SNCA* haplotypes for association with PD risk using logistic regression in LARGE-PD and a European-ancestry PD cohort from the International Parkinson Disease Genomics Consortium (IPDGC).

**Results:** The GWAS-significant PD PRS had an area under the receiver-operator curve (AUC) of 0.668 (95% CI: 0.640-0.695) and explained 2.2% of the phenotypic variance on the liability scale in LARGE-PD. The inclusion of external Peruvian data as controls mitigated this result, dropping the AUC 0.632 (95% CI: 0.607-0.657). In 1KGP Latinos, we found the PD PRS to exhibit a bias by ancestry. At the *SNCA* locus, haplotypes differ by ancestry. Ancestry-specific *SNCA* haplotypes were associated with PD status in both LARGE-PD and the IPDGC cohort (p-value < 0.05). Apart from rs356182, these haplotypes share as little as 14% of their variants.

**Conclusion:** The PD PRS has potential for PD risk prediction in Latinos, but variability caused by admixture patterns and bias in the PD PRS calculated using only European-ancestry data limits its utility. The inclusion of diverse subjects can help elucidate PD risk loci and improve risk prediction in non-European cohorts. In the case of the *SNCA* locus, by leveraging a Latino cohort, we provide orthogonal evidence for rs356182 causality.

## Introduction

Parkinson’s Disease (PD) is the fastest growing neurological disorder in the world, affecting more than six million individuals.^1^ Like all complex disorders, PD etiology is thought to be due to the combination of genetic and environmental risk factors, with the common variants of small effect comprising the major component of genetic risk factors.^2^ Genome-wide association studies (GWAS) have been used to identify genetic variants that modify disease risk and discover disease-related biological pathways. In PD, the largest GWAS effort to date is Nalls et al. 2019^3^ which features an impressive sample size of 37.7 thousand cases, 18.6 thousand proxy-cases, and 1.4 million controls. This study, however, only includes individuals with European ancestry, a common occurrence in GWAS data.^4^ Diversity in PD research is increasing: Foo et al. 2020 have conducted the largest study of PD patients with East Asian ancestry^5^ and our group has conducted the largest study of South American PD patients.^6^

Outside of risk variant and disease-gene discovery, a primary use of GWAS is to generate summary statistics for the purpose of risk prediction using polygenic risk scores (PRS). A PRS is the linear summation of disease risk variants weighted by their regression effect size and has been shown to improve disease risk prediction.^7^ The PRS model has been applied to an increasing number of diseases with the eventual goal of risk stratification followed by clinical interventions.^7^ In PD, Nalls et al. leverage their summary statistics in order to generate two PRS models, one incorporating only independent GWAS-significant variants and the other constructed using the full set of GWAS summary statistics, that demonstrate promise for PRS-based PD risk prediction.^3^

However, a major downside of PRS-risk prediction is the difficulty in transferring scores generated using GWAS from one population to another with a different ancestry background.^8,9^ It is thought that this lack of portability is primarily due to either differences in allele frequencies or linkage disequilibrium (LD) patterns, though differing gene-by-environment interactions could also be a factor.^4,10^ Ideally, representative population-matched GWAS data would be used to generate a PRS. However, since a PRS depends on accurate effect size estimates, very large sample sizes are needed to achieve adequate out-of-sample prediction.^11^ Due to the persistent lack of diversity in GWAS data, large sample sizes are typically only available for European or East Asian-ancestry subjects. This is a major challenge for the clinical implementation of PRS-based risk prediction and can exacerbate existing disparities.^10^

In PD, we also see the drop in performance when translating PRS across populations. Foo et al. applied a PRS based on the Nalls et al. GWAS-significant variants to PD patients from East Asia; the performance of the PRS lagged behind that of European cohorts, though this was remedied via the inclusion of Asian-specific data.^5^ Here, we construct a PRS using summary statistics from Nalls et al. 2019^3^ and tested it in our Latino case-control cohort from the Latin American Research Consortium on the Genetics of Parkinson’s Disease (LARGE-PD).^6,12^ In addition, we seek to characterize the distribution of PD risk alleles and the PD PRS across diverse global populations via the 1000 Genome Project^13^ and the Peruvian Genome Project.^14^ We also explore the haplotype structure of rs356182 near *SNCA*, a major component of the PD PRS and thought to be a key gene in PD etiology^15^, across ancestrally diverse populations.

## Methods

### LARGE-PD Cohort Description

1,504 LARGE-PD individuals from Uruguay, Peru, Chile, Brazil, and Colombia have genotype data available. Samples were genotyped using the Multi-ethnic genotyping array (MEGA) chip from Illumina.^16^ Genotyped subjects have a mean age of 59.3 (± 13.9) years; 44.3% are male and 55.7% female. Overall, the analysis dataset consists of 807 PD cases and 690 controls after quality control, with 1481 samples that feature complete age and sex records. PD patients were evaluated by a local movement disorder specialist using the UK PD Society Brain Bank clinical diagnostic criteria (UKPDSBB).^17^ Individuals who did not exhibit neurological symptoms were selected as controls. All participants provided written informed consent according to their respective locale’s national requirements. A complete description of LARGE-PD, including ancestry composition, ascertainment, quality control, and imputation, can be found in Loesch et al. 2020.^6^

### Additional PD Cohorts Description

See **supplementary table 1** for the description of all cohorts used in this study. For validating the PD PRS performance in Latinos, we utilized a cohort of Latinos provided by the International Parkinson Disease Genomics Consortium (IPDGC).^18^ These subjects were identified as Latinos based on principal components analysis and were excluded from the primary IPDGC GWAS. The first cohort, which will be referred to as NeuroX_C, consists of 448 subjects, with 223 controls and 225 cases (49.55% male, age statistics unavailable). Genotyping was done using neurogenerative disease-specific genotyping chips: 155 of the samples were genotyped using the NeuroX^19^ chip; the remainder were genotyped using the NeuroChip (NeuroC)^20^.

The IPDGC also provided 715 PD subjects and 1731 controls of European ancestry for our analysis of *SNCA* haplotypes, which we will refer to as IPDGC-EUR. All IPDGC-EUR subjects have undergone whole genome sequencing, have a mean age of 73 years (±18.4) and are 54.2% male.

### Non-PD Cohort Description

We utilized the unrelated subset of the high-coverage 1000 Genomes Project data generated by the New York Genome Center^21^ as references in our haplotype analysis and to estimate the PD PRS distribution across ancestral populations. We also utilized all sequenced Peruvian Genome Project^14^ samples as additional Native American references in our haplotype analysis.

We used 440 subjects over the age of 50 from a Peruvian tuberculosis (TB) cohort from Luo et al. to use as additional controls in order to evaluate the robustness of our PRS models.^22^ These samples have a mean age of 62.65 (SD: 9.13) years and are 46.4% male. They were genotyped using the Affymetrix LIMAArray, a custom array with 720,000 SNPs.^22^ For this study, we will refer to this cohort as Luo_TB.

### Imputation of Genotyped Samples

The NeuroX, NeuroC, and Luo_TB cohorts were filtered for 10% sample and site missingness, a Hardy-Weinberg exact (HWE) test p-value < 1×10^−6^, keeping only biallelic SNPs. We then imputed each cohort separately using the TOPMed Imputation Server hosted by the University of Michigan.^23^ The imputation pipeline employs Eagle2 for phasing, Minimac4 for imputation, and filters results by an R^2^ of 0.3. The TOPMed Imputation server has been shown to improve imputation for Latino populations and is currently the best publicly-available option.^23,24^

### PRS estimation and evaluation

We utilized summary statistics from Nalls et al. 2019^3^; we lifted the positions to hg38 using UCSC LiftOver utility. We first constructed a PRS using only the 90 independent genome-wide significant PD risk variants. After removing sites that were strand ambiguous (i.e. CG/AT), we calculated this PRS using R and PLINK 1.9^25^ with 77 variants. To protect against confounding, we repeated the overall analysis by resolving relative pairs via removing one sample as identified using the KING-robust software^26^ (testing 2^nd^ and 3^rd^ degree thresholds), by down-sampling the number of Peruvian PD cases, and by including additional external Peruvian controls in our dataset to ensure our results are not being driven by case-control imbalances. We also excluded subjects who were outliers by ancestry. For a previous study^6^, the ancestry proportions of LARGE-PD subjects were inferred by merging with 1000 Genomes Project (1KGP)^13^ subjects and using the software ADMIXTURE^27^ with a K of 5. Potential outliers were identified by selecting the subjects with three times the standard deviation greater or less than the mean of each of the 5 inferred clusters. Principal components were then re-computed with these subjects excluded.

We also assessed a PRS utilizing the full summary statistics from Nalls et al. 2019.^3^ For PRS models using the full summary statistics, we used PRSice-2^28^ to perform pruning and thresholding. We evaluated all estimated PRS models using R (see **supplementary methods**).

### Age at Onset Analysis of the GWAS-significant PRS

We assessed the impact of the PRS on the age at onset (AAO) of PD using a filtered dataset consisting of the unrelated LARGE-PD subjects as determined by KING^21^ with an AAO after the age of 18. We generated Kaplan–Meier curves of the GWAS-significant PRS stratified by quintile. For the event, we used the diagnosis of PD; for time to event, we used age of onset where available and age at analysis for controls and cases lacking age of onset data. We then performed a Cox regression analysis; we again stratified the PRS by quintile and adjusted for sex, the first 10 PCs, and recruitment site. We performed all analyses using the survival^26^ package in R. Plots were generated using the survminer package in R (https://CRAN.R-project.org/package=survminer).

### PRS validation

To validate the PD PRS performance, we repeated the GWAS-significant PRS analysis after incorporating external Peruvian controls from the Luo_TB cohort.^22^ After imputation, we calculated a PD PRS in all subjects from the TB cohort over the age of 50 using the same set of variants as before and combined this data with LARGE-PD. To calculate principal components, we merged imputed LARGE-PD and Luo_TB data, keeping the intersection of 7,122,988 variants with an imputation R^2^ of 0.9 and a MAF of 0.01 in both datasets. We then performed two rounds of pruning using PLINK’s indep-pairwise algorithm with parameters of 50 SNPs per window, a step of 5 SNPs, and an R^2^ of 0.2, leaving us with 341,969 variants for the estimation of PCs and a kinship matrix using PC-AiR^29^ and PC-Relate^30^. To evaluate the quality of the merger, we calculated GC Lambda from a GWAS on 766,828 SNPs obtained from the first LD pruning step. We performed the GWAS using a logistic mixed model implemented by the GENESIS R package^31^, adjusting for age, sex, the first 10 PCs, and the genetic relationship matrix obtained from PC-Relate. We evaluated the PRS in the same manner as previously described, though in this case the full model included age, sex, the first 10 PCs, recruitment site, and study.

We also tested both the GWAS-significant and the full summary statistics PRS in an independent cohort of 448 samples (NEUROX_C). Out of the 1040 variants used in the PRS constructed with the full PD summary statistics, 950 were imputed with a minimum imputation R^2^ of 0.8 in NeuroC samples, but only 651 were imputed with a minimum imputation R^2^ of 0.8 in the intersection of the NeuroC and NeuroX samples.

### PD PRS distribution in LARGE-PD

We visualized the PRS distribution in LARGE-PD and the external Peruvian controls using R. For clustering by PC, we constructed a distance matrix by taking the Euclidean distance of the first two PCs after scaling. Then, we performed k-means clustering via the kmeans function in R, using the Hartigan-Wong algorithm, 5 centers, a maximum of 100 iterations, and 10 random sets. The ancestry of each cluster was then inferred via the mean ancestry proportions of subjects within the cluster obtained using the ADMIXTURE^27^ software as described above.

### Characterization of GWAS-significant loci across diverse populations

We characterized the distribution of the PD PRS across global populations using the high coverage data from the 1000 Genomes Project (1KGP)^13^ data generated by the New York Genome Center. We calculated the PD PRS as described above. Differences in PD PRS distribution across 1000 Genomes super-populations (AFR, AMR, EAS, EUR, SAS) were assessed using the Wilcoxon rank-sum test by using the EUR populations as a reference. To assess differences in risk allele frequencies across super-populations, we created a contingency table for each non-European super-population based on direction of effect and allele frequency. For direction of effect, we counted variants with a positive beta coefficient; for allele frequency, we counted variants with a higher frequency in the given super-population compared to EUR populations. We then tested each contingency table using the Chi-Square test with a single degree of freedom. We also explored the relationship of admixture with the PD PRS in 1KGP by utilizing the ancestry proportions estimated with ADMIXTURE as described above and obtaining correlations between each ancestry proportions and the PD PRS using Pearson’s method.

### *Haplotype Analysis of SNCA* region

We generated a joint dataset including all phase III 1KGP samples, sequenced Peruvian Genome Project data, imputed LARGE-PD data, and IPDGC-EUR sequence data, including only the intersection of variants across all datasets. The merged dataset was then jointly phased using Beagle 5.0 on default settings.^32^ We again utilized PLINK’s haplotype block procedure to estimate haplotype blocks in this region. We then extracted a region corresponding to the LARGE-PD Peruvian haplotype block containing rs356182.

Using R, we parsed the phased VCF file, keeping SNPs with a MAF of 5% or higher. We generated frequency counts for each unique haplotype and binned these counts according to PD study, case-control status, 1KGP super-population, and rs356182 allele status. Haplotype similarities were assessed by determining the number of variants shared between each haplotype. We then constructed a haplotype network using POPArt^33^ and the TCS method^34^; to simplify the network and prevent a PD recruitment bias, we removed haplotypes that did not achieve 1% frequency in 1KGP Data.

For LARGE-PD and IPDGC data, we tested haplotypes with a frequency higher than 1% in each respective cohort for association with PD risk. With LARGE-PD, we utilized the unrelated subset and adjusted for age, sex, recruitment site, and the first 10 PCs. With the IPDGC data, we also adjusted for age, sex, cohort, and the first five PCs. Multiple testing correction was applied by adjusting for the number of haplotypes tested in each cohort. Haplotypes with a p-value less than 0.05 were then tested using a likelihood ratio test to assess whether the addition of the haplotype demonstrates significant improvement over a model including all covariates and the rs356182 genotype status of each subject.

## Results

### PD PRS in the LARGE-PD cohort

We found the GWAS-significant PD PRS to be highly associated with PD status (p-value=1.91×10^−18^) and it explained 2.2% of trait variance on the liability scale (see **Table 1**) even though it was derived from European GWAS data. When stratifying the PRS by quintile, the highest quintile had an odds ratio of 5.38 (95% CI: 3.78-7.67) when compared to the lowest quartile (**Figure 1A**). The PRS efficiently separated cases and controls (**supplementary figure 1A**). Using only the PRS to predict PD risk, the area under the receiver-operator curve (AUC) was 0.668 (95% CI: 0.640-0.695; see **Figure 1B, supplementary figure 1B**), with a balanced accuracy of 61.7%, a sensitivity of 71.3% and a specificity of 52.1 % (see **Table 1**). The addition of the GWAS-significant PD PRS to a model including all covariates improved the AUC by 4.3% over the base model without the PRS; this improvement was statistically significant with a p-value of 1.03×10^−6^ (Delong’s test). We also constructed a PRS using the full summary statistics (PRS-full) generated by Nalls et al. and the same parameters utilized by Nalls et al. in their study. The PRS resulted in the inclusion of 1040 variants and had an overall AUC of 0.676 (95% CI: 0.649-0.704**; Table 1**), with a balanced accuracy of 61.5%, a sensitivity of 69.8% and a specificity of 53.1%. The AUCs of the GWAS-significant and full summary stat models were not significantly different (p-value = 0.44, Delong’s test).

**Table 1:** PRS results.

| COHORT | SUBSET | PRS TYPE | PVALUE | R2 | PSEUDO R2 | AUC (95% CI) | ACC (BAL) | SPEC | SENS |
| --- | --- | --- | --- | --- | --- | --- | --- | --- | --- |
| LARGE-PD | ALL | GWAS SIGNIF | 1.91x10<sup>-18</sup> | 0.022 | 0.058 | 0.668 (0.640-0.695) | 0.625 (0.617) | 0.521 | 0.713 |
|  | UNREL – 2<sup>nd</sup> Degree |  | 1.68x10<sup>-18</sup> | 0.023 | 0.059 | 0.668 (0.640-0.696) | 0.627 (0.619) | 0.518 | 0.721 |
|  | UNREL – 3<sup>rd</sup> Degree |  | 1.08x10<sup>-17</sup> | 0.022 | 0.057 | 0.666 (0.638-0.694) | 0.631 (0.622) | 0.520 | 0.725 |
|  | OUTLIERS |  | 3.36x10<sup>-18</sup> | 0.021 | 0.055 | 0.666 (0.638-0.694) | 0.626 (0.619) | 0.527 | 0.710 |
|  | DOWN SAMPLED |  | 6.32x10<sup>-16</sup> | 0.022 | 0.057 | 0.664 (0.634-0.694) | 0.632 (0.626) | 0.535 | 0.717 |
|  | PERU ONLY |  | 8.60x10<sup>-11</sup> | 0.028 | 0.068 | 0.675 (0.635-0.716) | 0.670 (0.618) | 0.379 | 0.857 |
|  | PERU EXCL. |  | 1.48x10<sup>-08</sup> | 0.016 | 0.044 | 0.629 (0.589-0.668) | 0.599 (0.594) | 0.490 | 0.697 |
|  | ALL | FULL SUM STATS | 2.37x10<sup>-22</sup> | 0.028 | 0.072 | 0.676 (0.649-0.704) | 0.621 (0.615) | 0.531 | 0.698 |
| LARGE-PD + EXTERNAL CONTROLS | ALL | GWAS SIGNIF | 2.18x10<sup>-18</sup> | 0.015 | 0.038 | 0.632 (0.607-0.657) | 0.620 (0.577) | 0.319 | 0.834 |
|  | UNREL |  | 2.31x10<sup>-19</sup> | 0.015 | 0.039 | 0.635 (0.608-0.660) | 0.623 (0.580) | 0.326 | 0.833 |
|  | PERU ONLY |  | 6.25x10<sup>-11</sup> | 0.012 | 0.029 | 0.645 (0.612-0.677) | 0.639 (0.557) | 0.229 | 0.885 |
| NEUROX +<br>NEUROC | ALL | GWAS<br>SIGNIF | 4.19x10<sup>-05</sup> | NA | NA | 0.655<br>(0.604-0.705) | 0.596<br>(0.596) | 0.583 | 0.609 |
|  |  | FULL<br>SUM<br>STATS | 5.16x10<sup>-06</sup> | NA | NA | 0.662<br>(0.612-0.712) | 0.607<br>(0.607) | 0.601 | 0.613 |

COHORT: cohort label. SUBSET: subpopulation label from cohort. PRS TYPE: type of model used (either GWAS significant SNPs or the full summary stats). PVALUE: p-value of the PRS in a logistic regression model. R2: variance explained on the liability scale. PSEUDO R2: Nagelkerke's Pseudo R<sup>2</sup>. AUC (95% CI): area under the receiver-operator curve and 95% confidence intervals. ACC (BAL): accuracy and balanced accuracy of PRS alone. SPEC: specificity of the PRS alone. SENS: sensitivity of the PRS alone.

**Figure 1:**
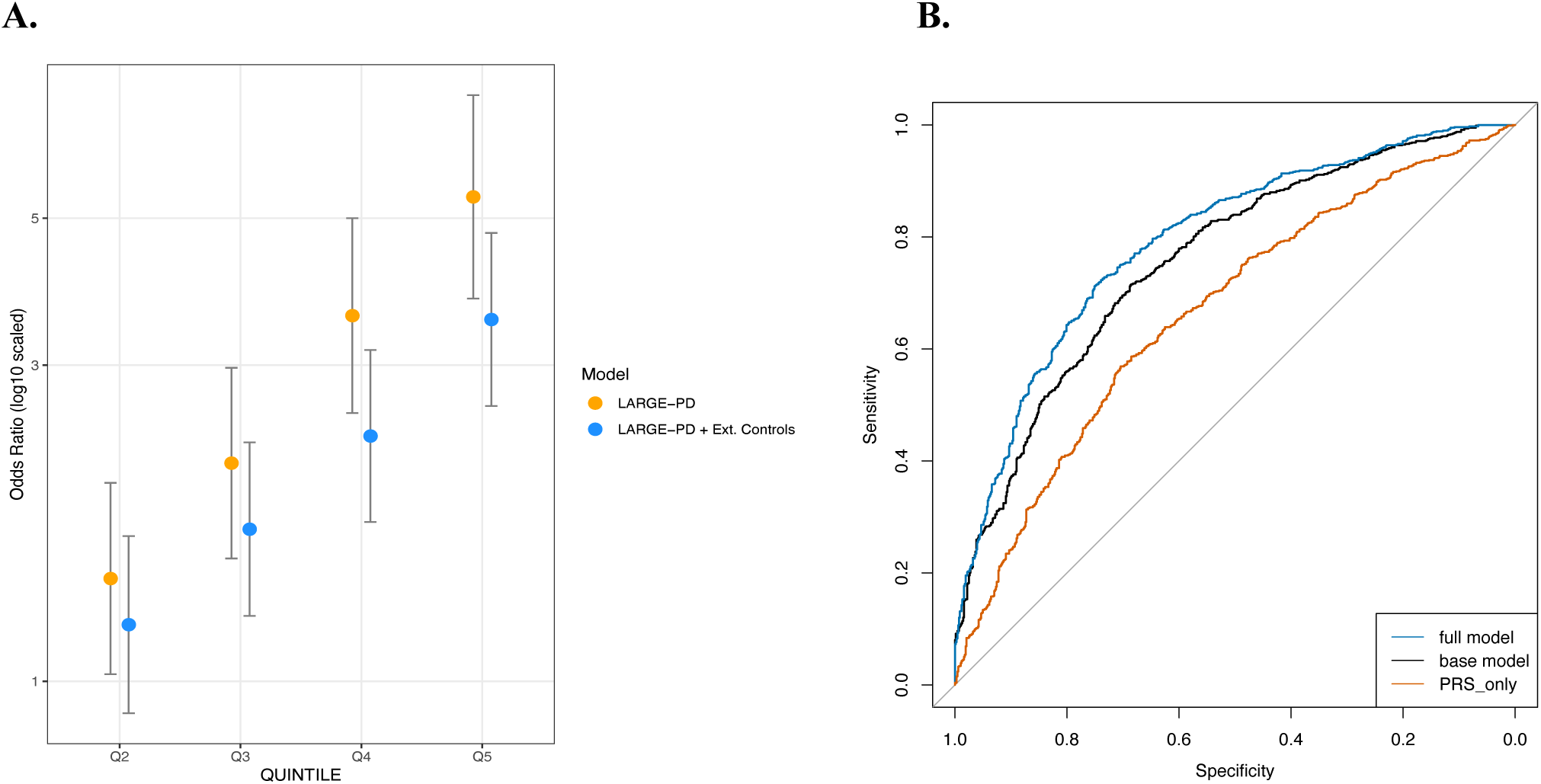
PD PRS prediction in LARGE-PD. **A:** Odds ratios of LARGE-PD subjects with a PD PRS in quintiles 2 through 5 compared to quintile 1 (orange) and odds ratios of LARGE-PD subjects plus the addition of external Peruvian controls (blue). **B:** Receiver-operator curve (ROC) of full model including covariates (age, sex, PCs 1-10, recruitment site) and the GWAS-significant PD PRS (blue), ROC of base model including only covariates (black), and ROC of model including only the GWAS-significant PD PRS (orange). The addition of the GWAS-significant PRS to create the full model improved the AUC by 4.3% over the base model without the PRS (p-value of 1.03×10^−6^, Delong’s test). All models shown in 1B include only data from LARGE-PD.

### Age at Onset Analysis

To evaluate the impact of the PD PRS on disease onset, we generated Kaplan-Meier curves and performed Cox’s proportional hazard regression using the age at analysis for controls and age at onset for cases (see **methods**). We stratified the GWAS-significant PRS by quintile and found that the age at onset (AAO) decreased when comparing the highest quintile to the lowest (see **Figure 2**). In our Cox regression model, the highest quintile had a hazard ratio (HR) of 2.29 (95% CI: 1.79-2.93; p-value: 3.41×10^−11^). We also repeated the analysis using only cases, with the PRS still being significantly associated with AAO, though the effect is attenuated (HR: 1.45, 95% CI: 1.14 - 1.86, p-value: 0.003; see **Table 2**).

**Table 2:** Cox regression results.

| QUINTILE | HR | 95% CI | PVALUE |
| --- | --- | --- | --- |
| CASES ONLY |  |  |  |
| 2 | 1.10 | 0.832- 1.44 | 0.515 |
| 3 | 1.14 | 0.879 - 1.48 | 0.324 |
| 4 | 1.13 | 0.878 - 1.46 | 0.341 |
| 5 | 1.45 | 1.17 - 1.91 | 0.003 |
| CASES + CONTROLS |  |  |  |
| 2 | 1.28 | 0.984 - 1.68 | 0.096 |
| 3 | 1.43 | 1.11 - 1.85 | 0.007 |
| 4 | 1.60 | 1.24 - 2.06 | $2.55 \times 10^{-04}$ |
| 5 | 2.29 | 1.79 - 2.93 | $3.41 \times 10^{-11}$ |

QUINTILE: quintile of the PD PRS. HR: hazard ratio. 95% CI: 95% confidence interval of the hazard ratio. PVALUE: p-value of the hazard ratio.

**Figure 2:**
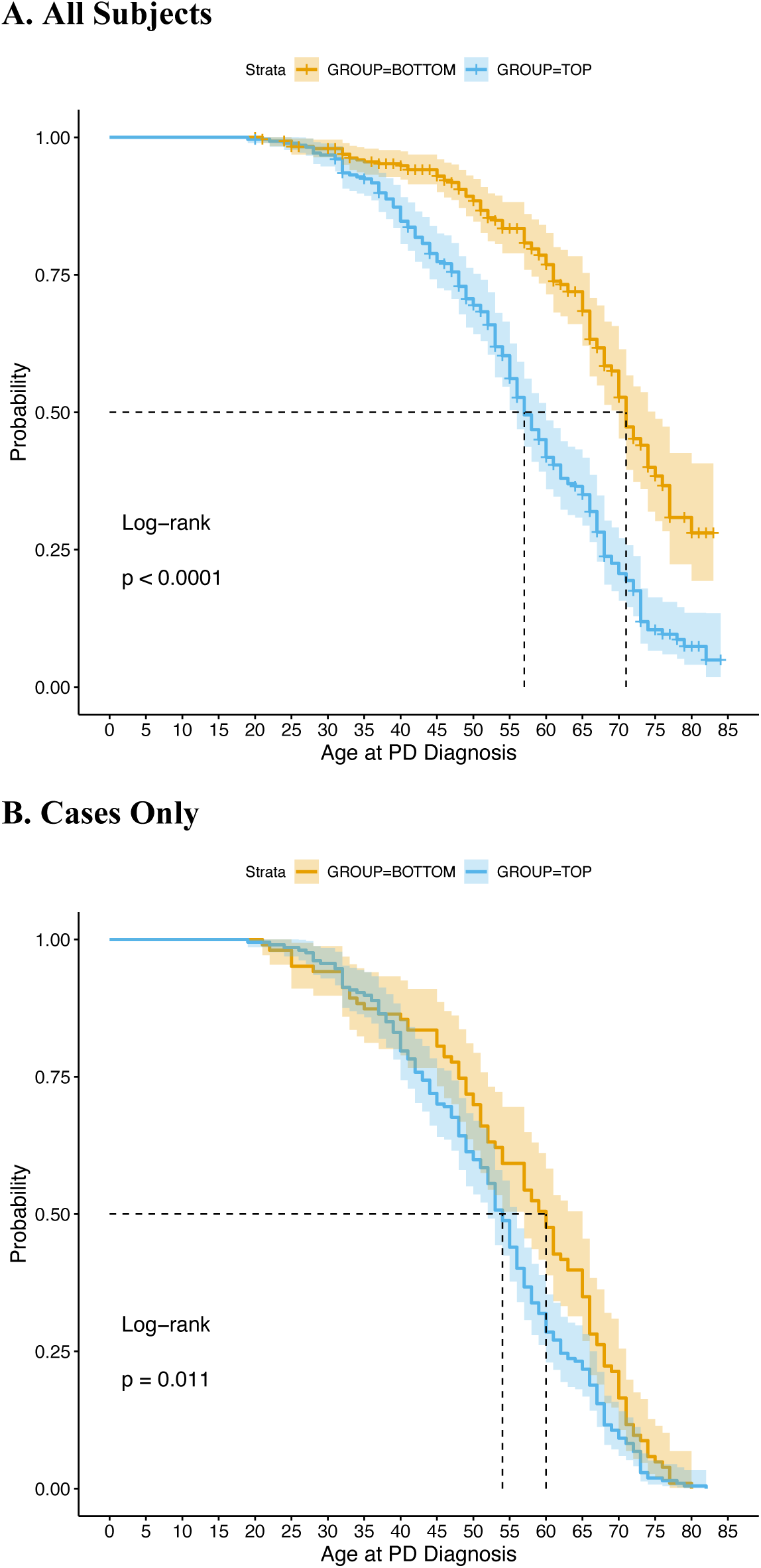
PD PRS Survival Analysis. Kaplan-Meier curves of age at PD onset stratified by GWAS-significant PRS quintile in all subjects (A) and in cases only (B) generated using the survival package in R. For controls, age at analysis was used. Cases with an age at onset less than 18 years were excluded.

### PD PRS validation

The predictive performance of the PD PRS was remarkable as LARGE-PD is a Latino cohort with a mean European ancestry of only 47%.^6^ In LARGE-PD, the AUC of the GWAS-significant PRS exceeded the 0.655 AUC reported by Nalls et al. in their out-of-sample prediction for a European dataset.^3^ Also contrary to expectations the performance of the GWAS-significant PRS was driven by LARGE-PD participants from Lima, Peru (see **supplementary figure 1B; Table 1**), who are predominantly of Native American ancestry. This result was robust to both removing close relatives and down-sampling Peruvian PD cases (see **Table 1**). We also evaluated whether uncorrected population stratification caused by the presence of outliers by ancestry could have contributed to the observed results. In LARGE-PD, removal of outliers by ancestry resulted in the exclusion of 37 subjects of primarily African ancestry and 12 subjects with East Asian ancestry (see **methods**). The performance of the PD PRS in LARGE-PD was robust to the removal of potential outliers (see **Table 1**) and the AUC was not significantly different from the AUC obtained in the full LARGE-PD cohort (p-value = 0.9561, Delong’s test). In addition, the beta coefficients estimated for the Nalls et al. GWAS variants using the outlier-removed LARGE-PD subset were 99.7% correlated with those estimated in the full LARGE-PD cohort^6^, suggesting that the inclusion of these ancestral outliers did not impact these analyses.

To further evaluate this result, we employed both additional external controls and an independent Latino cohort. When adding 440 additional Peruvian controls from an external study^22^, the AUC of the GWAS-significant PRS dropped to 0.632 (95% CI: 0.607-0.657), with a balanced accuracy of 57.7%, a specificity of 31.9% and a sensitivity of 83.4% (see **Table 1, supplementary figure 1B**). Furthermore, the variance explained on the liability scale was only 1.5%, though this could be partially attributed to the choice of covariates and the use of external controls that were not screened for PD. Though the AUC is substantially lower with the inclusion of the external controls, the AUCs of models with and without the external controls were not significantly different (p-value = 0.08, Delong’s test). To ensure that this result is not driven by population stratification introduced by the inclusion of external controls, we performed a GWAS with 766,828 SNPs (see **methods**). The GC lambda of this GWAS was 0.995, suggesting that population stratification is not a factor here. In addition, the lead SNP is a variant in *SNCA*, as would be expected given prior PD GWAS results.^3,6^

To validate the GWAS-significant PRS and the PRS-full, we tested both models in a cohort of 448 Latinos provided by the IPDGC. The GWAS-significant model had an AUC of 0.665 with a balanced accuracy of 59.6%, a sensitivity of 61.3% and a specificity of 60.0% (see **Table 1**). For the PRS-full, only 651 of the variants were imputed at a sufficient level across both genotyping chips used (see **methods**). Even using only 62.6% of the SNPs, the AUC was 0.662 (95% CI: 0.612-0.712) (**supplementary figure 2**).

### PD PRS distribution in LARGE-PD

The variability of models using the PD PRS can be visualized by examining the PD PRS distribution by country (see **Figure 3 A and B**). Excluding Chilean subjects due to sample size, subjects from Peru had the highest mean PRS (mean [SD]: 0.18 [0.55]) while samples from Colombia had the lowest (mean [SD]: −0.04[0.62]). This was not attributable to case-control ratio, as the mean PRS was not significantly correlated with the proportion of cases (p-value = 0.75). However, the first four PCs were all significantly correlated with the PD PRS (p-value < 0.05). When clustering samples by PC, the PD PRS distributions reflect the ancestral compositions of the clusters, with inferred African clusters shifted to the left of zero and inferred Native American clusters shifted to the right (see **Figure 3 C and D**). These biases where the entire distribution is shifted makes interpreting the PRS performance difficult. In addition, due to variability in local ancestry patterns, it is possible for variability to exist in the PD PRS distribution within one geographic area. For example, the PD PRS distribution varied among our samples from Lima, Puno, and the external controls from Peru-wide recruitments (see **Figure 3 E and F**).

**Figure 3:**
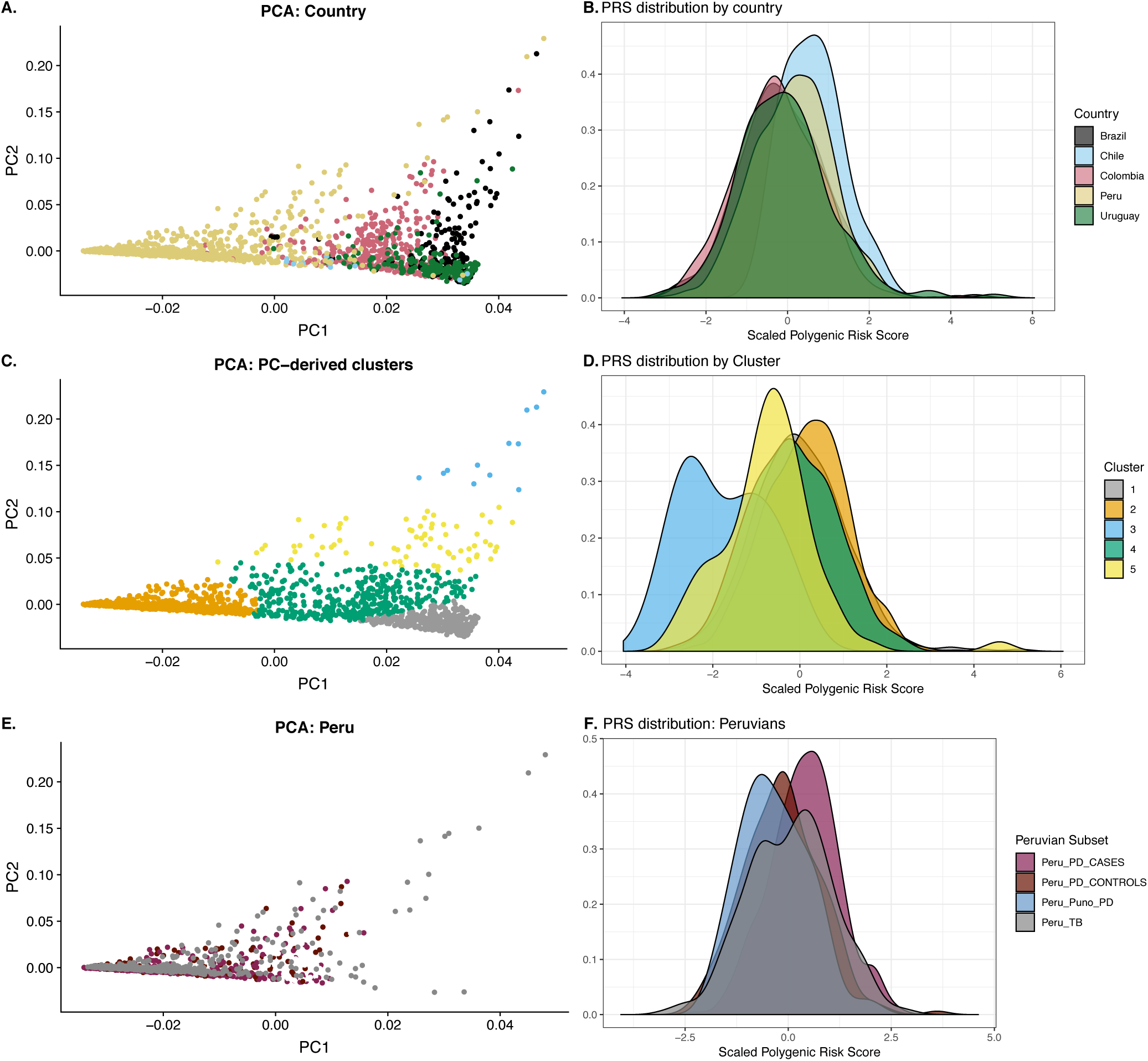
PRS distribution in LARGE-PD and external controls. Principal components (PCs) of LARGE-PD plus external controls and density plots of the PD PRS distribution. **A:** Plot of PC 1 versus PC 2 colored by country of origin. **B:** PRS distribution colored by country of origin. **C:** Plot of PC 1 versus PC2 colored by PC-derived clusters using k-means clustering. **D:** Distribution of the PD PRS colored by PC-derived clusters. We used ancestry proportions estimated by ADMIXURE to characterize clusters. We observed that the African-ancestry cluster (blue) was shifted to the left, the European-ancestry cluster (silver) was centered on zero, and the Native American cluster (orange) was shifted to the right. **E:** PCA plot of subjects from Peru. Subjects are classified as being cases, controls, from Puno (all controls), or external Peruvian controls. **F:** PD PRS distribution in Peru.

### GWAS-significant PD risk variants across diverse human populations

To highlight the variability in PD risk loci across diverse ancestral populations, we calculated a GWAS-significant PD PRS for every individual in phase III of the 1000 Genomes Project^13^ (1KGP) using the high-coverage data generated by the New York Genome Center.^21^ The PD PRS was lowest in African populations and highest in East Asian populations (see **supplementary figure 3**). For each non-European population, we assessed the difference in PD PRS distribution compared to Europeans using the Wilcoxon Rank-sum test (see **supplementary table 2**). The PD PRS distribution significantly differed from European-ancestry samples for every other global population.

Differences in the PRS distribution is likely being mediated by population-specific differences in allele frequencies. When plotting European PD risk allele frequencies by African allele frequencies and labeling the points by the variant’s estimated odds ratio, variants conferring positive disease risk are demonstrably lower in frequency in African populations as has been previously noted^35^ (see **supplementary figure 4)**. By constructing contingency tables of allele frequency differences (higher vs. lower than EUR allele frequencies) and effect size direction (risk vs. protective), we found that the count of risk alleles with lower allele frequencies to be significant in African populations (p-value= 0.001, Chi-square test with 1 df).

The 1KGP includes Latinos from Peru, Mexico, Puerto Rico, and Colombia. These individuals are admixed with varying contributions from African, European, and Native American ancestral populations. We estimated ancestry proportions using ADMIXTURE and the full 1KGP dataset (see **supplementary figure 5**). In Latinos, the PD PRS is positively correlated with inferred Native American ancestry (Pearson’s R: 0.19, p-value: 0.0004) and negatively correlated with both European (Pearson’s R: −0.11, p-value: 0.03) and African ancestry (Pearson’s R: −0.30, p-value: 1.4×10^−8^).

We also estimated a polygenic risk score using the 71 risk SNPs in common across every available Peruvian cohort (see **supplementary table 3**). LARGE-PD Peruvian controls have a lower mean PD PRS compared to Peruvian subjects from 1KGP (0.54 versus 0.58) or the PGP (0.68). When stratifying the PGP by sub-population, we observed a fair amount of heterogeneity, with the PRS ranging from a mean of 0.58 (the Uros) to 0.88 (the Chopccas).

### Haplotype Analysis for rs356182 locus

The SNP rs356182, at the *SNCA* locus, is the lead variant in the large European-ancestry meta-analyses^3^ and has the largest expected effect size among common PD Risk variants, thus playing a large role in the PD PRS. Furthermore, it was the only GWAS-significant variant in both LARGE-PD and a Hispanic/Latino 23andMe replication cohort.^6^ To analyze haplotypes at this locus, we merged imputed LARGE-PD data with 1KGP, Peruvian Genome Project, and IPDGC-EUR sequence data. Using PLINK, we estimated haplotype blocks in the *SNCA* region that contain rs356182 for select populations in our dataset (**supplementary table 4**). For most global populations, the rs356182 haplotype block is small due to recombination. Within the PD cohorts, the largest haplotype blocks were consistently found in Peruvian populations. We extracted the 33.6 kilobase (kb) region corresponding to the block found in the Peruvian subset of LARGE-PD. In this region, we categorized the haplotypes based on their rs356182 allele status (see **Figure 4A**). The same A-allele haplotype is the most common in all out-of-Africa populations in our joint dataset, while the G-allele haplotype appears to be more population specific. Interestingly, the most common East Asian G-allele haplotype shares more variants with the most common A-allele haplotype than does the most common European G-allele haplotype (**supplementary figure 6**). We constructed a haplotype network using haplotypes with a minimum frequency of 1% in the 1KGP and the TCS method as implemented by PopArt^33^ (see **Figure 5**). The most common G-allele haplotype in East Asians (hap9) and the most common G-allele haplotype in Europeans (hap1) are separated by a number of intermediary haplotypes, again demonstrating their independence.

**Figure 4:**
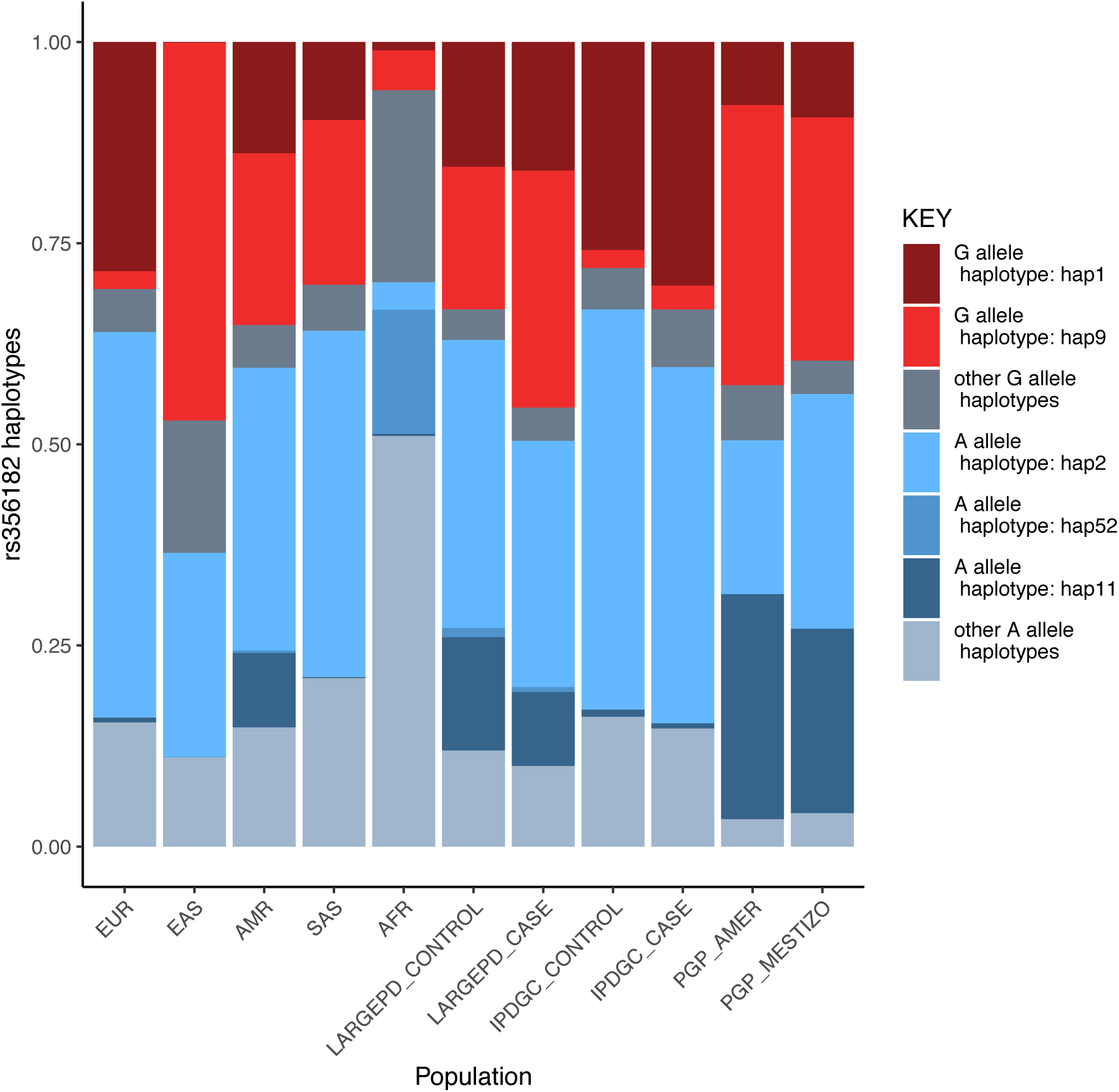
rs356182 Haplotypes by population. Haplotypes of the *SNCA* locus centered on rs356182 consisting of phased genotypes from the 1KGP, LARGE-PD, and an IPDGC PD cohort of European descent. Haplotypes shown here are the most common haplotypes by 1000 Genomes population and PD case-control status. Note the same A haplotype is shared across populations, but the G haplotype exhibits greater population specificity.

**Figure 5:**
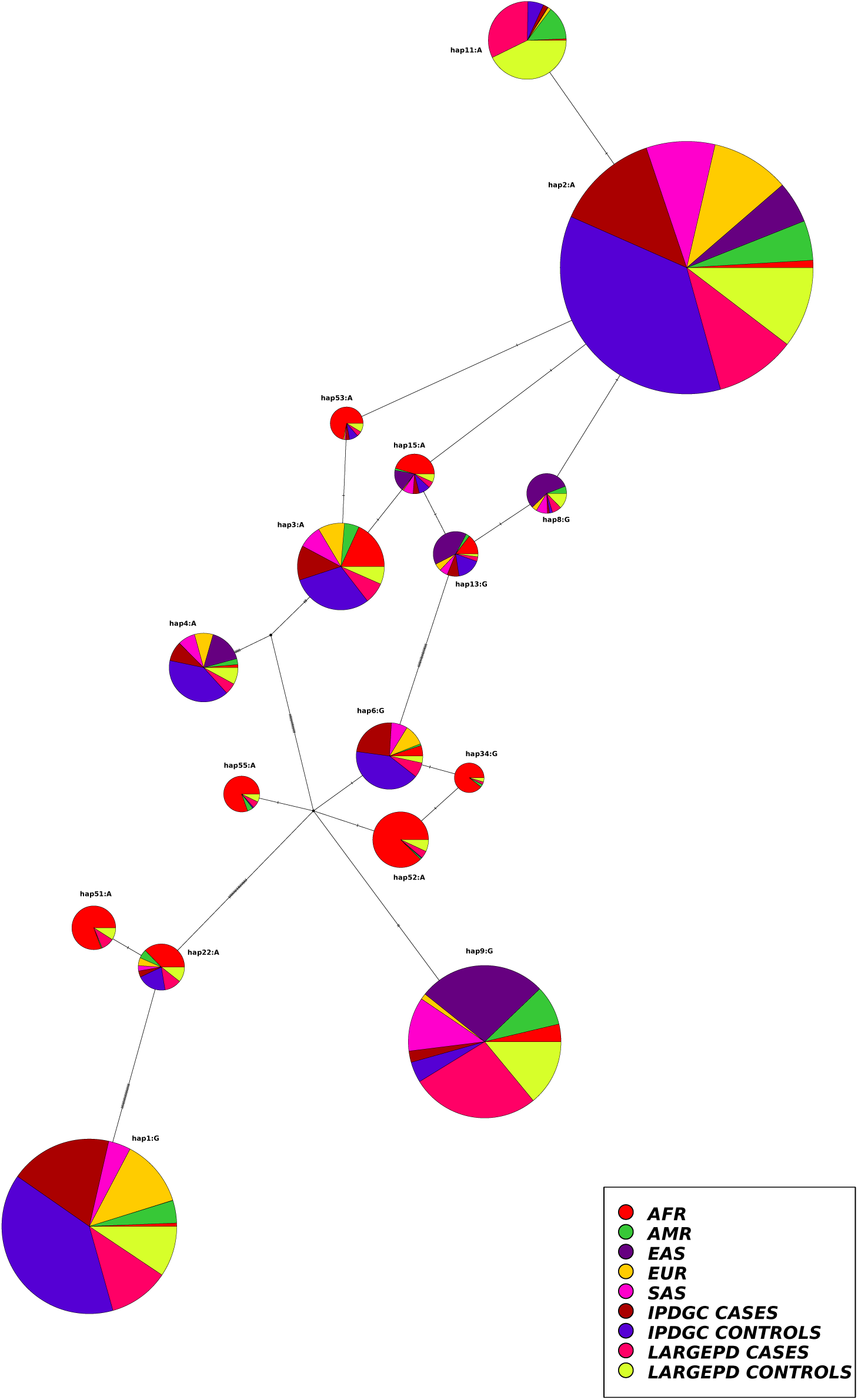
Haplotype Network of rs356182 haplotypes. Haplotype network of haplotypes from 1KGP, LARGE-PD, and an IPDGC PD cohort of European descent as constructed using the TCS network algorithm and POPArt. Note the presence of two distinct G-allele haplotype clusters which were separated by A-allele haplotypes.

We selected rs356182 haplotypes with a frequency of 1% in LARGE-PD or the IPDGC-EUR cohort and tested them for association with PD using logistic regression (see **methods, supplementary tables 5** and **6**). The direction of effect for seven out of eight haplotypes tested in LARGE-PD and six out of seven haplotypes tested in IPDGC were concordant with rs356182 allele status. In LARGE-PD, three haplotypes were nominally associated with PD status: hap6 (p-value = 0.006), hap9 (p-value = 4.47×10^−^ 6), and hap11 (p-value = 6.04×10^−9^); all three haplotypes remained significant after adjusting for multiple tests (adjusted p-value < 0.05). In IPDGC-EUR, two haplotypes were nominally associated with PD: hap1 (p-value = 0.01) and hap2 (p-value = 1.75×10^−4^). After correcting for multiple testing, hap2 remained statistically significant. We then evaluated whether the addition of haplotype information demonstrated significant improvement over a model that included rs356182 allele status using a likelihood ratio test. In LARGE-PD, the hap6 and hap11 were nominally significant (p-value 0.039 and 2.65×10^−5^, respectively) while hap9 was not nominally significant (p-value 0.076). After correcting for multiple testing, only hap9 remained statistically significant (adjusted p-value < 0.05). In the IPDGC-EUR, hap1 and hap2 were not nominally significant (p-values 0.29 and 0.15, respectively).

## Discussion

Polygenic risk prediction has the potential to identify individuals at higher risk of developing disease who could benefit from interventions and increased monitoring. However, PRS predictive performance depends on GWAS with large sample sizes which are generally only available in European-ancestry cohorts and, to a lesser extent, East Asian ancestry cohorts. The performance of PRS derived from such datasets suffers when applied to individuals from a different ancestral background, leading to inaccurate or even biased estimates of disease risk.^8–10^ In addition, a PRS derived from one ancestry can exhibit shifts in distribution across ancestries that are not necessarily concordant with population-level disease risk. Interpreting and ultimately rectifying these shifts is an area of ongoing research. For PD, the sample size is now sufficiently large that future clinical application of the PD PRS is plausible, though the European bias could potentially limit its utility. In a large East Asian cohort, Foo et al. found that the GWAS-significant PD PRS using only European data performed notably worse (AUC 0.602) than the PRS that incorporated population-specific data (AUC 0.631).^5^ However, even when including East Asian data, this still performed worse than a European-ancestry GWAS-significant PRS in a European cohort (AUC 0.655).^3^ Bandres-Ciga et al. tested a machine learning approach and a PRS, both constructed using the full set of GWAS summary statistics, in a Spanish PD cohort. The machine learning approach had an AUC of 0.6205 while the PRS model had an AUC that was approximately 1% lower.^36^ This was notably worse than the performance of a full summary statistics PRS tested in Nalls et al.^3^ (AUC 0.692), indicating that there is also a potential Northern European bias.

In LARGE-PD, a Latino PD cohort, we found that the PD PRS constructed using GWAS-significant data performed surprisingly well at face value, with an AUC of 0.668 (95% CI: 0.640-0.695), outpacing the AUC of the European and East Asian cohorts when utilizing the same set of GWAS-significant PD summary statistics (0.655 and 0.602 AUC, respectively). This result runs counter to the bulk of the PRS literature; predictive performance should be worse when applying a PRS across ancestries. While it is possible that these GWAS-significant variants might play an outsized role in the etiology of PD in Latinos, a more parsimonious explanation is that bias in the GWAS summary statistics, together with the complex composition of the LARGE-PD cohort, contribute to the performance we observed. Using data from both LARGE-PD and external cohorts, we do find evidence that the PD PRS exhibits population-wide shifts in distribution. In 1KGP subjects, we found that the PD PRS distribution significantly differs from that of Europeans in all other populations (p-value < 0.05, Wilcoxon). This is likely due to differences in allele frequencies. In both LARGE-PD and 1KGP Latinos, we found that the PD PRS exhibits a bias by ancestry where individuals with high Native American ancestry tended to have a higher PD PRS, while individuals with high African ancestry were more likely to have a lower PD PRS (see **Figure 3, supplementary figure 5**).

In LARGE-PD, PD cases had a higher mean Native American ancestry than controls which could be contributing to the surprisingly strong performance of the PD PRS as measured by AUC due to population stratification aligning with the by-ancestry bias of the PD PRS. In the case of Peruvian subjects, the AUC remained high even when we down-sampled Peruvian cases and when we fit models with only Peruvian subjects (see **Table 1**). However, when we included external Peruvian subjects as controls, we saw a reduction in the AUC to 0.632 (95% CI: 0.607-0.657). We calculated a PD PRS for every Peruvian subject available to us and found that Peruvian LARGE-PD controls from Lima and Puno have a lower mean PRS than any other subpopulation. This suggests that LARGE-PD controls have been sampled from the lower end of the PD PRS distribution in Peru, either purely due to chance or because they belong to a subpopulation with a lower frequency of some of the PD risk alleles used in the PRS. Together, these results suggest that the PD PRS is impacted by population history, heterogeneity exists even within a single country, and that PRS performance metrics without the use of covariates were likely inflated.

Despite the challenges, the use of a PD PRS for risk prediction in Latinos certainly has potential. In all scenarios we tested, the PD PRS achieved a degree of separation between cases and controls (see **Table 1**). In addition, PD cases with a PD PRS in the highest quintile had a hazard ratio of 1.45 (95% CI: 1.17 - 1.91) compared to PD cases in the lowest quintile, demonstrating that the PD PRS contributed to the modification of disease course. Due to the bias in the PD PRS distribution, care needs to be taken when interpreting results and the inclusion of covariates are necessary to mitigate confounding. As demonstrated by LARGE-PD, the admixture patterns in a cohort can have a strong impact on the PRS performance. Before it can be used in the clinic, the challenges of translating the PD PRS across populations will need to be addressed through the inclusion of diverse GWAS data and through improved methods development, ideally in an admixture-aware manner.

As the GWAS variant with the largest expected effect size among common variants (frequency > 5%), rs356182 in SNCA contributes to a significant proportion of the variance explained in the PD PRS. Shifts in rs356182 frequency or exclusion of this SNP due to poor genotyping can have a large impact on the predictive accuracy of the PD PRS. Consequently, we conducted a haplotype analysis at the SNCA locus centered on rs356182. In general, haplotype blocks were small in most populations, with the exception of Peruvian and East Asian cohorts, and rs356182 was not well tagged (defined as an r2 > 0.8) in any non-PD cohort due to recombination. An examination of the most frequently seen haplotypes in this region, though, reveals global patterns. Nearly every population shares the same common A-allele haplotype, while the most common G-allele haplotype in European-ancestry individuals differs from East Asian, South Asian, and Native American individuals (see **Figure 5**). In LARGE-PD, the non-European G-allele haplotype (hap9) was robustly associated with PD status, while in a European-ancestry cohort, the European G-allele haplotype was nominally significant. These two haplotypes only share 14% of their alleles (see **supplementary figure 6**) and are likely independently derived from the African A haplotype, which points towards rs356182 as a causal variant contributing risk, as has previously been suggested through functional data.^15^ Furthermore, in both cohorts, the direction of effect of each haplotype was overwhelmingly concordant with its rs356182 allele status and the inclusion of four of five nominally significant haplotypes did not significantly improve models with rs356182 genotype status after correcting for multiple tests. Interestingly, the addition of the hap11 haplotype significantly improved the rs356182 model even after correcting for multiple tests. This haplotype only differs from hap2, the globally common A-allele haplotype in this region, by a single variant (rs12505231). This variant has not been characterized in the PD literature and the score predicted by the Combined Annotation and Depletion algorithm^37^ (CADD) does not suggest a functional role (PHRED-scaled CADD = 1.65), though this variant is more common in African and Latin American populations than European populations (1KGP). While rs12505231 could potentially be playing a hitherto undescribed role in PD etiology, it is also possible that this haplotype includes a relevant variant not captured in the 30 kb window examined for this study. Additional studies of the *SNCA* region in diverse populations will be critical for identifying and characterizing functional variation.

Our study was limited by sample size, particularly on the country or sub-population level. The predictive performance of the PD PRS could differ in larger Latino populations, particularly if the ancestral composition differs from that of LARGE-PD. In addition, our use of external samples as controls could introduce a degree of error due to not being explicitly screened for PD status, though we should be able to assume a population-level prevalence rate of 1%. Our estimation of haplotype blocks depended on the variants that remained in the intersection of the merged datasets, so subtle differences can result from the selection of datasets used in the merger. Despite these limitations, we show both the potential and shortcomings of utilizing a European-ancestry PD PRS in non-European cohorts and highlight the bias in the PD PRS by ancestry. We also provide orthogonal evidence that rs356182 is a causal variant, rather than simply tagging a causal variant, and again highlight the value of including diverse data in the analysis of PD risk loci.

## Data Availability

All data produced in the present study are available upon reasonable request to the authors

## Acknowledgements

We thank all of the individuals who participated in LARGE-PD. We also want to thank all the support staff at the different Latin American sites for their efforts and support in this project. We also thank members of the International Parkinson Disease Genomics Consortium (IPDGC) for their contributions of both data and expertise to this project. In particular, we want to thank Cornelis Blauwendraat and Mike A. Nalls for their insights.

## Data Availability

Code used for this project can be accessed at www.github.com/dloesch/LARGEPD_PRS. 1000 Genomes Project sequence data can be found at https://www.internationalgenome.org/. International Parkinson’s Disease Genomics Consortium (IPDGC) data is available here https://pdgenetics.org/resources and additional inquiries regarding IPDGC data can be made at https://pdgenetics.org/contact. Peruvian Genome Project data is available through the European Genome-Phenome Archive (EGA): https://ega-archive.org/datasets/EGAD00001007082. Data from Luo et al. is available on the database of Genotypes and Phenotypes (dbGaP) with accession number phs002025.v1.p1. LARGE-PD genotype data will be uploaded to dbGaP for recruitment sites that have completed the dbGaP certification process. Summary statistics for the full LARGE-PD cohort are currently available in the PD GWAS browser: https://pdgenetics.shinyapps.io/GWASBrowser/.

