## Supplemental Methods and Data for "Polygenic risk prediction and *SNCA* haplotype analysis in a Latino Parkinson’s disease cohort"

*PRS estimation and evaluation*

Custom R code was used to generate the GWAS-significant PD PRS. For the PD PRS using the full GWAS summary statistics, PRSice-2 was utilized.[1] PRSice-2 iterates over a range of p-values to select the parameters that explain the most trait variance. PRSice-2 calculates the observed variance using Nagelkerke's pseudo $R^2$, converting to the liability scale using a given prevalence, in this case 0.5%. It is recommended to utilize a LD-reference panel that matches the population used to generate the summary statistics, so we utilized 500 European samples from the 1000 Genomes Project[2] as the LD reference panel.

Both the custom code and PRSice-2 calculated the PRS using the following equation:

$$PRS_i = \sum_{j=1}^{M} \frac{\beta_j X_{ij}}{M}$$

where the PRS of individual *i* is calculated by taking the summation of the individual's genotype (0,1,2) of SNP *j* weighted by its corresponding effect size estimated in a large-scale GWAS and averaged by the *M,* number of SNPs used in the PRS construction.

We followed the same testing paradigm for all PRS models via 10-fold cross validation procedure and a logistic regression framework. The full logistic model includes age, sex, recruitment site, the first 10 PCs, and the PRS; the base model includes all terms except the PRS. We calculated Nagelkerke's pseudo $R^2$ using the DescTools[3] R package and estimated the variance explained on the liability scale using the method described by Lee et al.[4] We obtained the observed $R^2$ by subtracting the base model $R^2$ from the full model $R^2$; we then converted this to the liability scale using a prevalence of 0.5% and the proportion of cases in LARGE-PD.[4,5] We determined the area under the receiver operator-curve (AUC) using the pROC package[6] in R and predictions generated from the 10 folds using both the full model and the PRS alone. Statistical significance between the AUCs obtained in different models was determined using Delong's test via the pROC package. We also obtained p-values and the Pseudo $R^2$ via the mean of the 10 folds.

### Supplemental Figures

#### Supp. Figure 1: Performance of PD PRS in LARGE-PD

**A.** **B.**

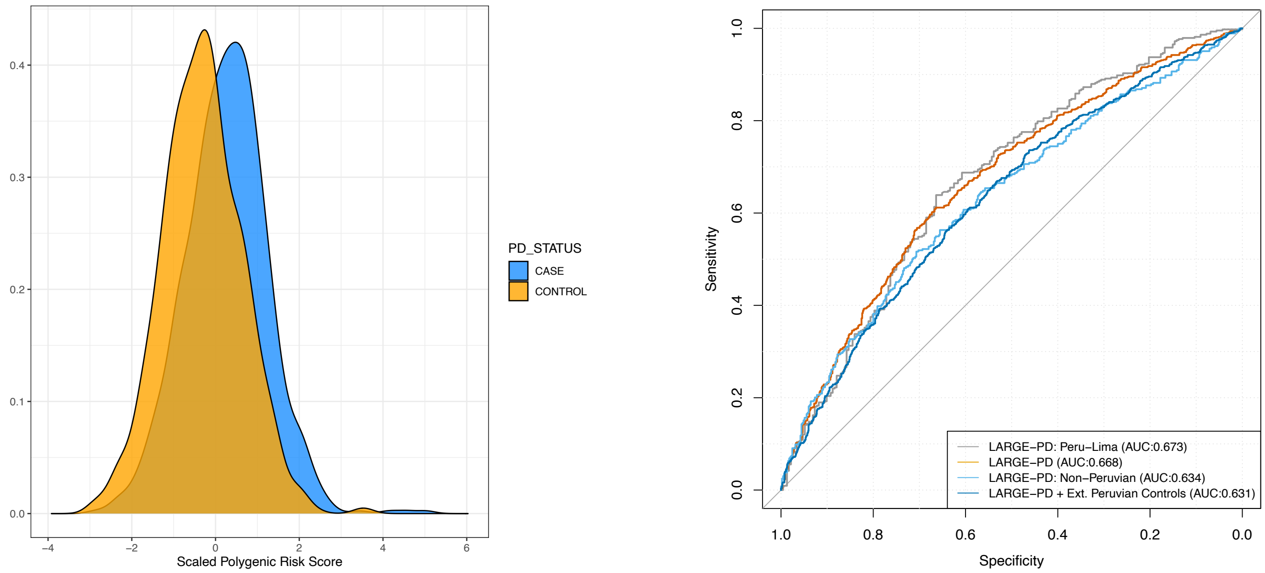


**A:** Distribution of the PD PRS constructed using GWAS-significant variants in LARGE-PD cases versus controls. **B:** Plots of the receiver operator curve (ROC) when predicting PD status using the PD PRS alone for Peruvian LARGE-PD subjects (silver), all of LARGE-PD (red), non-Peruvian LARGE-PD subjects (light blue), and LARGE-PD plus 440 external Peruvian controls (light blue). ROC curves were generated using the pROC package in R and the PD PRS consists of only independent GWAS-significant variants.

**Supp. Figure 2: AUC in NEUROX_C cohort of Latino subjects.**

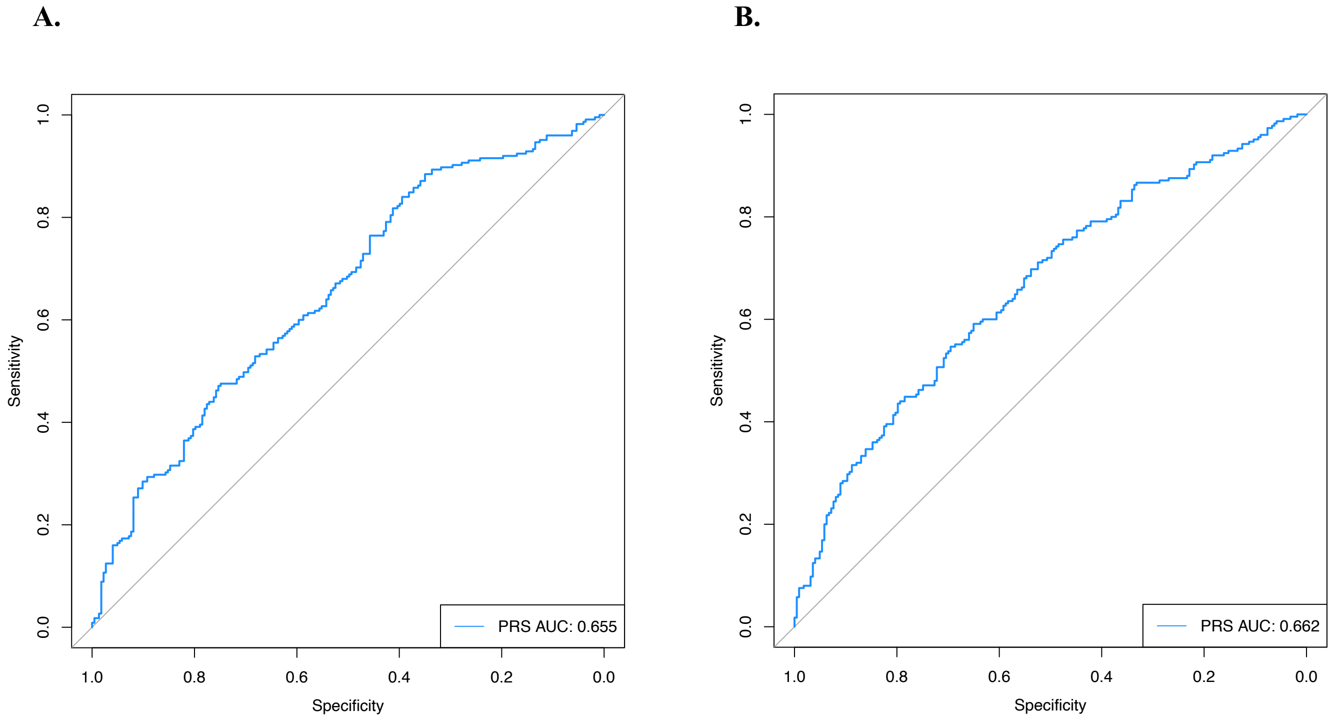


**A:** Receiver-operator curve (ROC) of the PD PRS constructed using only GWAS-significant variants in the NeuroX_C cohort supplied by the IPDGC. **B:** ROC of the PD PRS constructed using the full PD GWAS summary statistics from Nalls et al. 2019.

**Supp. Figure 3: PRS distribution by 1000 Genomes Project Super-Population**

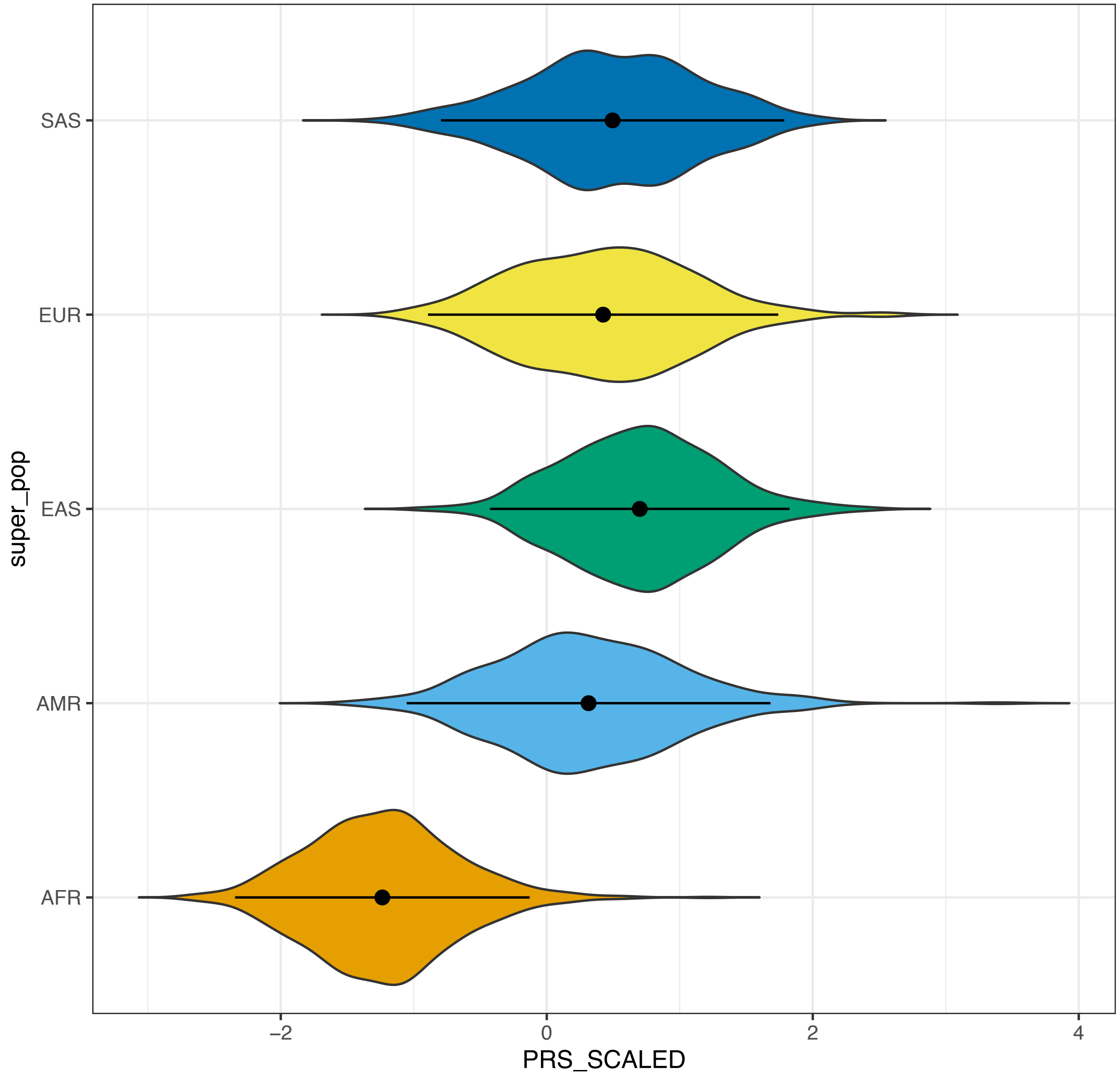


Distribution in the 1000 Genomes Project (1KGP) of the scaled PD PRS constructed using GWAS-significant variants in the 1KGP-defined super-populations of AFR (African), AMR (Admixed American i.e., Latinos), SAS (South Asian), EAS (East Asian), and EUR (European).

**Supp. Figure 4: PD risk allele frequencies in African versus European populations**

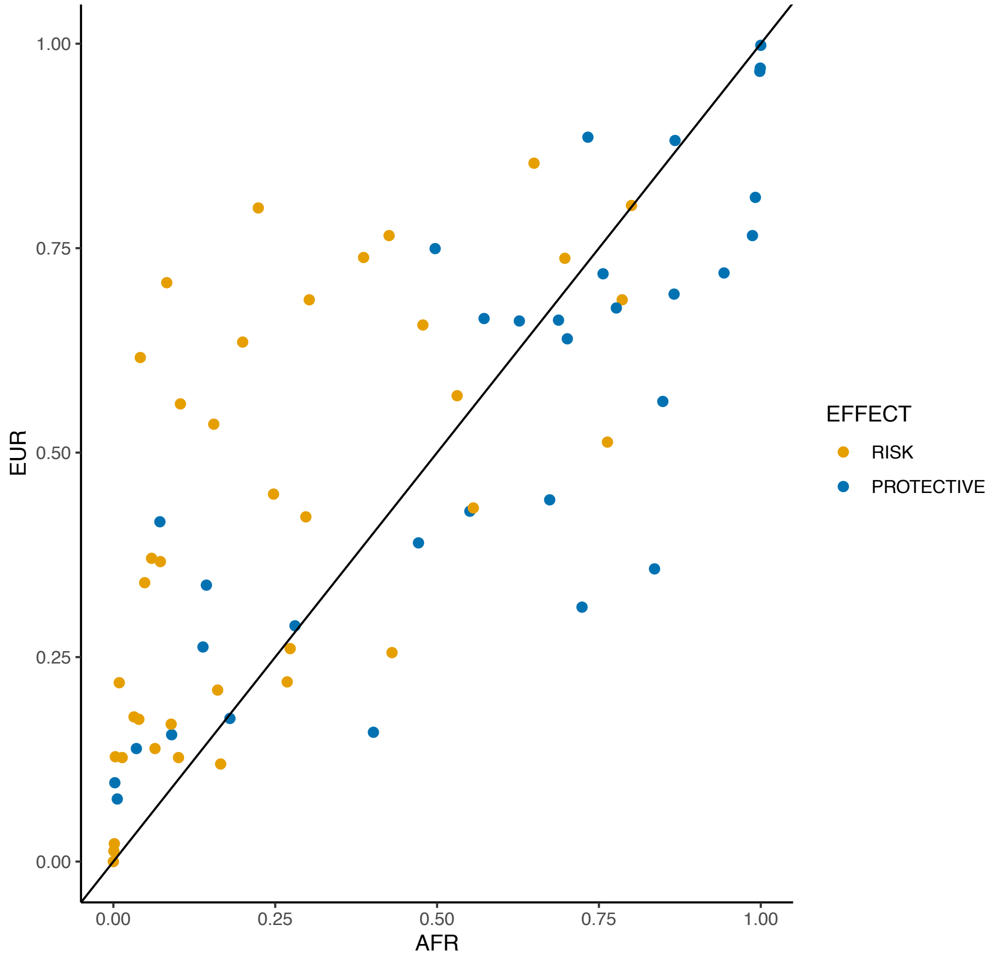


Scatter plot of PD risk allele frequencies in African populations (X-axis) versus European populations (y-axis) from the 1000 Genomes Project. Variants with a positive direction of effect were labeled as risk (orange) while variants with a negative direction of effect were labeled protective (blue). Variants above the black line are higher in frequency in European populations, while variants below the black line are more frequent in African populations. Note the preponderance of risk variants with a higher frequency in European populations.

### Supp. Figure 5: PD PRS by Ancestry in 1KGP Latinos

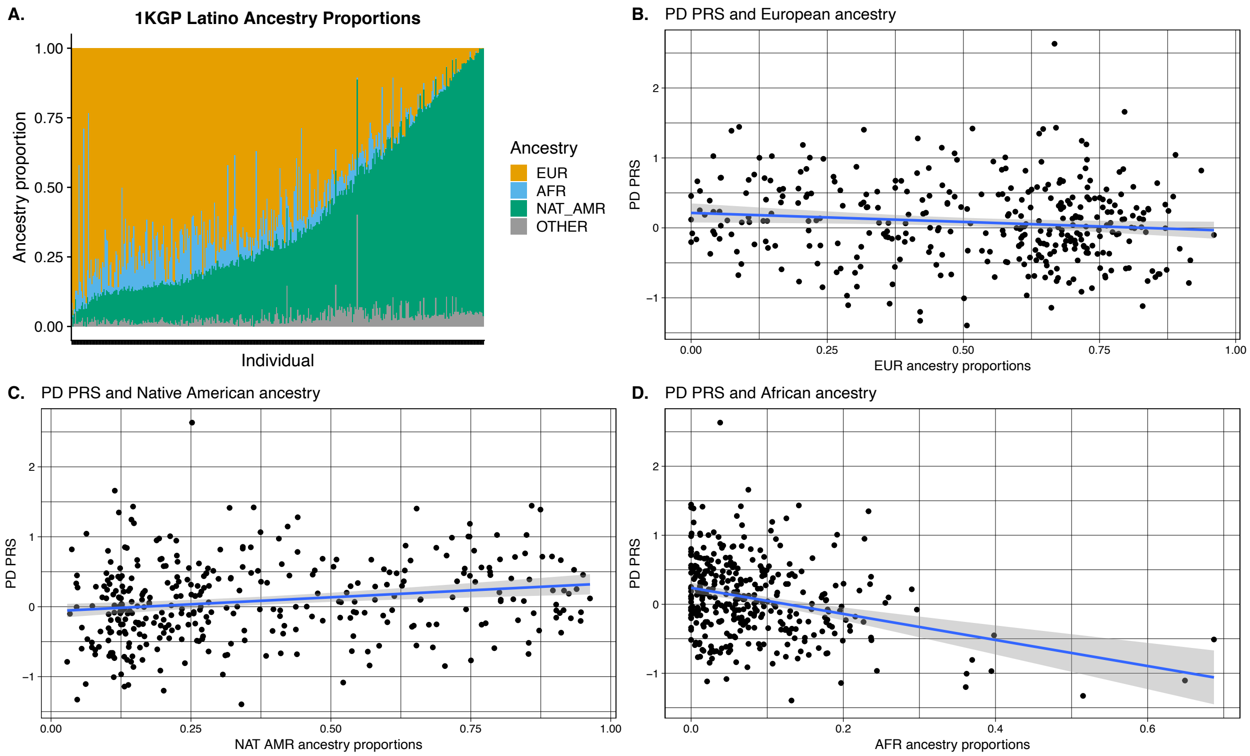


**A:** Ancestry proportions of 1KGP Latinos as estimated by ADMIXTURE. **B:** Scatterplot of PD PRS versus European ancestry proportion with the regression line in blue (95% confidence interval in grey). **C:** Scatterplot of PD PRS versus Native American ancestry proportion with the regression line in blue (95% confidence interval in grey). **D:** Scatterplot of PD PRS versus African ancestry proportion with the regression line in blue (95% confidence interval in grey).

**Supp. Figure 6: Heatmap of shared alleles between common rs356182 haplotypes**

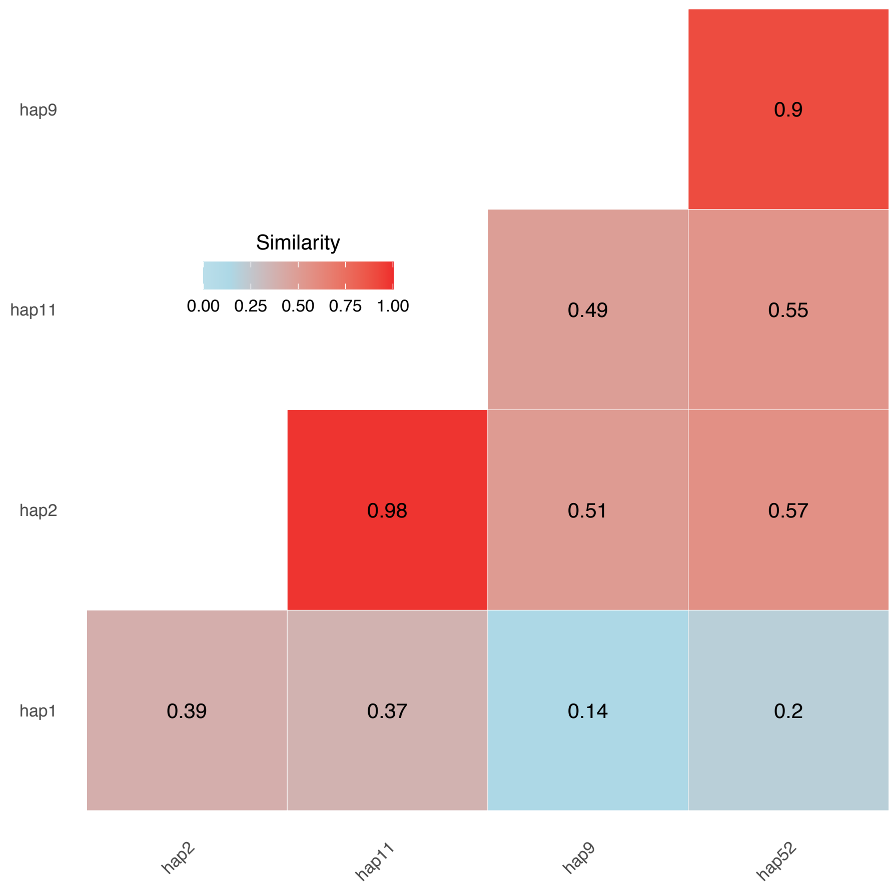


Heatmap of the proportion of shared alleles between common rs356182 haplotypes. For example, the haplotypes hap1 and hap9 share 0.14 or 14% of their alleles, including rs356182. Haplotypes were extracted from a 33.6 kb region from the merged 1000 Genomes Project, Peruvian Genome Project, LARGE-PD, and IPDGC data.

**Supplemental Tables**

**Supp. Table 1: Cohort Description**

| Cohort | N (N Cases) | Mean (SD) Age | Sex | Data Type | Recruitment Country |
|---|---|---|---|---|---|
| LARGE-PD | 1497 (807) | 59.3 (13.9) | 44.3% male | Genotyped + imputed | Peru, Brazil, Colombia, Chile, Uruguay |
| External Controls (Luo_TB) | 440 (0) | 62.65 (9.13) | 46.4% male | Genotyped + imputed | Peru |
| IPDGC-Latino (NeuroX_C) | 448 (225) | Not known | 49.55% male | Genotyped + imputed | USA (Latino/Hispanic) |
| IPDGC-European | 2446 (715) | 73 (18.4) | 54.2% male | Sequenced | USA, UK (European-ancestry) |
| Peruvian Genome Project (PGP) | 150 (0) | NA | NA | Sequenced | Peru |
| 1000 Genomes Project (1KGP) | 2504 (0) | NA | 49.2% male | Sequenced | Multiple |

N (N Cases): Number of subjects (Number of cases). Mean (SD) Age: Mean and SD of age at analysis. Sex: Proportion of the cohort that is male. Data Type: Indicates type of genetic data, i.e., sequenced or genotyped. Recruitment Country: Primary country for cohort recruitment.

**Supp. Table 2: PRS by 1000 Genomes Super-Population**

| POP | MEAN | SD | PVAL |
|---|---|---|---|
| **EUR** | 0.17 | 0.55 | NA |
| **EAS** | 0.4 | 0.47 | $3.49x10^{-12}$ |
| **AMR** | 0.08 | 0.57 | 0.02 |
| **SAS** | 0.23 | 0.54 | 0.05 |
| **AFR** | -1.21 | 0.46 | $2.02x10^{-169}$ |

POP: 1KGP super-population label. MEAN: mean PRS. SD: standard deviation of the PRS. PVAL: p-value of the Wilcoxon test with European subjects as reference.

**Supp. Table 3: PD PRS in Peruvian Populations**

| GROUP | N | MEAN | SD | P-VALUE |
|---|---|---|---|---|
| **PEL** | 85 | 0.581 | 0.562 | NA |
| **PERUVIAN_CASES** | 437 | 0.865 | 0.508 | $2.03x10^{-5}$ |
| **PERUVIAN_CONTROLS** | 233 | 0.542 | 0.519 | 0.6058 |
| **PUNO** | 45 | 0.500 | 0.509 | 0.3679 |
| **TB_ALL** | 4009 | 0.641 | 0.497 | 0.2764 |
| **TB_CONTROLS** | 440 | 0.661 | 0.510 | 0.2169 |
| **PGP-ALL** | 150 | 0.679 | 0.462 | 0.1658 |
| **PGP-CHOPCCAS** | 30 | 0.880 | 0.499 | 0.02204 |
| **PGP-CUSCO** | 16 | 0.624 | 0.450 | 0.7273 |
| **PGP-IQUITOS** | 16 | 0.633 | 0.470 | 0.561 |
| **PGP-MATZES** | 12 | 0.802 | 0.385 | 0.1348 |
| **PGP-MOCHES** | 30 | 0.599 | 0.411 | 0.8635 |
| **PGP-TRUJILLO** | 16 | 0.638 | 0.609 | 0.5928 |
| **PGP-UROS** | 30 | 0.585 | 0.378 | 0.9467 |

GROUP: group label. N: number of subjects. MEAN: mean of raw PRS. SD: standard deviation of PRS. P-VALUE: p-value of PRS using the Wilcoxon rank sum test and PEL subjects as a reference.

**Supp. table 4: SNCA haplotype blocks in select populations**

| POPULATION | COHORT | SIZE (KB) | NSNP |
|---|---|---|---|
| **GIH** | 1KGP-SAS | 0.029 | 2 |
| **STU** | 1KGP-SAS | 0.029 | 2 |
| **FIN** | 1KGP-EUR | 0.029 | 2 |
| **CEU** | 1KGP-EUR | 0 | 0 |
| **CLM** | 1KGP-AMR | 0.029 | 2 |
| **PEL** | 1KGP-AMR | 0.303 | 3 |
| **ASW** | 1KGP-AFR | 0 | 0 |
| **YRI** | 1KGP-AFR | 0 | 0 |
| **CHB** | 1KGP-EAS | 68.573 | 87 |
| **JPT** | 1KGP-EAS | 12.424 | 18 |
| **CHOPCCAS** | PGP | 0.029 | 2 |
| **CUSCO** | PGP | 0 | 0 |
| **IQUITOS** | PGP | 96.637 | 131 |
| **MATZES** | PGP | 108.851 | 187 |
| **MOCHES** | PGP | 0 | 0 |
| **TRUJILLO** | PGP | 0 | 0 |
| **UROS** | PGP | 2.155 | 4 |
| **IPDGC_CONTROL** | IPDGC | 19.504 | 16 |
| **IPDGC_CASE** | IPDGC | 0.029 | 2 |
| **LARGEPD_CONTROL** | LARGEPD | 0.303 | 3 |
| **LARGEPD_CASE** | LARGEPD | 42.802 | 40 |
| **Chile** | LARGEPD | 0 | 0 |
| **Brazil_PortoAlegre** | LARGEPD | 0 | 0 |
| **Brazil_SaoPaolo** | LARGEPD | 0 | 0 |
| **Brazil_RibeiraoPreto** | LARGEPD | 0.029 | 2 |
| **Colombia_Bogota** | LARGEPD | 0 | 0 |
| **Colombia_Medellin** | LARGEPD | 0.327 | 3 |
| **Peru (Lima)** | LARGEPD | 33.628 | 57 |
| **Peru_Puno** | LARGEPD | 108.622 | 131 |
| **Uruguay** | LARGEPD | 0.029 | 2 |

POPULATION : population label. COHORT: cohort label. SIZE: haplotype size in kilobases. NSNP: number of SNPs in the haplotype.

**Supp. table 5: IPDGC rs356182 haplotype analysis**

| hapID | ALLELE | FREQ | FREQ CASES | FREQ CONT | BETA (SE) | PVAL (ADJ) | P_LRT | CONC |
|---|---|---|---|---|---|---|---|---|
| **hap1** | G | 0.28 | 0.3 | 0.26 | 0.36 (0.1) | 0.01 (0.089) | 0.29 | TRUE |
| **hap10** | A | 0.01 | 0.01 | 0.01 | 1.56 (1.1) | 0.14 (1) | . | FALSE |
| **hap2** | A | 0.48 | 0.44 | 0.5 | -0.5 (0.1) | $1.75 \times 10^{-4}$ (0.001) | 0.15 | TRUE |
| **hap3** | A | 0.05 | 0.05 | 0.05 | -0.1 (0.3) | 0.71 (1) | . | TRUE |
| **hap4** | A | 0.04 | 0.02 | 0.04 | -0.6 (0.4) | 0.13 (0.91) | . | TRUE |
| **hap6** | G | 0.04 | 0.06 | 0.04 | 0.42 (0.3) | 0.2 (1) | . | TRUE |
| **hap9** | G | 0.03 | 0.03 | 0.02 | 0.68 (0.5) | 0.15 (1) | . | TRUE |

hapID: haplotype ID. ALLELE: A or G allele for rs356182. G is the risk allele. FREQ: frequency of the haplotype. FREQ CASES: frequency of the haplotype in cases. FREQ CONT: frequency of the haplotype in controls. BETA (SE): effect size estimated in logistic regression model and standard error of the beta. PVAL (ADJ): p-value of the beta and adjusted p-value after correcting for number of haplotypes tested. P_LRT: p-value from likelihood ratio test evaluating whether inclusion of haplotype information improves models with rs356182 genotype status. CONC: Concordance of the direction of effect with rs356182 allele status.

**Supplemental table 6: LARGE-PD rs356182 haplotype analysis**

| hapID | ALLELE | FREQ | FREQ CASES | FREQ CONT | BETA (SE) | PVAL (ADJ) | P_LRT (ADJ) | CONC |
|---|---|---|---|---|---|---|---|---|
| **hap1** | G | 0.16 | 0.16 | 0.16 | 0.12 (0.12) | 0.287 (1) | . | TRUE |
| **hap11** | A | 0.11 | 0.09 | 0.14 | -0.81 (0.14) | $6.04x10^{-9}$ ($4.83x10^{-8}$) | $2.65x10^{-5}$ ($7.96x10^{-5}$) | TRUE |
| **hap2** | A | 0.33 | 0.3 | 0.36 | -0.14 (0.09) | 0.118 (0.944) | . | TRUE |
| **hap22** | A | 0.01 | 0.01 | 0.01 | 0.32 (0.4) | 0.418 (1) | . | FALSE |
| **hap3** | A | 0.03 | 0.03 | 0.03 | 0.42 (0.26) | 0.112 (0.898) | . | TRUE |
| **hap4** | A | 0.02 | 0.01 | 0.02 | -0.58 (0.36) | 0.106 (0.852) | . | TRUE |
| **hap6** | G | 0.01 | 0.02 | 0.01 | 1.18 (0.43) | 0.006 (0.049) | 0.039 (0.117) | TRUE |
| **hap9** | G | 0.24 | 0.3 | 0.17 | 0.5 (0.11) | $4.47x10^{-6}$ ($3.59x10^{-5}$) | 0.076 (0.228) | TRUE |

hapID: haplotype ID. ALLELE: A or G allele for rs356182. G is the risk allele. FREQ: frequency of the haplotype. FREQ CASES: frequency of the haplotype in cases. FREQ CONT: frequency of the haplotype in controls. BETA (SE): effect size estimated in logistic regression model and standard error of the beta. PVAL (ADJ): p-value of the beta and adjusted p-value after correcting for number of haplotypes tested. P_LRT (ADJ): p-value from likelihood ratio test evaluating whether inclusion of haplotype information improves models with rs356182 genotype status and p-value adjusted for multiple testing. CONC: Concordance of the direction of effect with rs356182 allele status.
